# Recovery from catatonia follows a hierarchical precision order

**DOI:** 10.64898/2026.08.13.26360194

**Authors:** Hiroki Saito, Yuichi Takizawa, Issei Murata, Amane Tateno, Jakob Theorell, Ryosuke Arakawa, Mikael Tiger

## Abstract

When benzodiazepines fail in catatonia, clinicians know what to escalate to but not what to stabilize first. We asked a narrower question that clinical records can address: does recovery follow a fixed order? We defined five capacities in plain clinical terms and mapped them onto precision domains of hierarchical inference. The domains are theory-derived clinical abstractions, not latent variables estimated from these data. In 25 consecutive inpatients managed without electroconvulsive therapy, an order fixed before coding held without inversion in every patient, and held again when a hypothesis-blind co-author re-rated all milestones according to a pre-registered protocol. In five patients the sequence was completed while hallucinations and delusions persisted, so the order does not follow the resolution of psychotic symptoms. Bush–Francis scores fell from 26.3 to 2.0. This retrospective study shows that the order held, not why. Prospective studies, preferably with blinded daily rating, are needed to corroborate the order.

## Introduction

Catatonia is a distinctive neuropsychiatric syndrome characterized by abnormalities of motor behaviour, volition, speech, and autonomic regulation.^1,2^ Contemporary diagnostic systems, including the Diagnostic and Statistical Manual of Mental Disorders, Fifth Edition, Text Revision (DSM-5-TR) and the International Classification of Diseases, 11th Revision (ICD-11), formalize it through observable criteria, improving diagnostic reliability.^3,4^ Yet despite this phenomenological definition, catatonia remains mechanistically unresolved, and its marked heterogeneity is still managed largely through empirical rather than principled approaches, a clinically consequential limitation, given its substantial morbidity and potential mortality if not promptly recognized and treated.^5^Benzodiazepines and electroconvulsive therapy (ECT) are the cornerstones of treatment and are often effective,^1,2^ yet a substantial minority of patients respond incompletely to first-line benzodiazepines,^6,7^ and some show paradoxical worsening or excessive behavioural suppression.^8^ Reported response to ECT is high, between 59% and 100% across studies, including after benzodiazepine non-response,^9^ so the difficulty is rarely the absence of an effective escalation but the absence of a rule for which capacity to stabilize first, particularly where ECT is unavailable, contraindicated or delayed. Escalation can attenuate overt motor manifestations without restoring coherent agency or functional reintegration, and these outcomes are often regarded as idiosyncratic.^9,7^ A central limitation is that catatonia is defined almost exclusively at the level of surface phenomena; DSM-5-TR phenomenology, though clinically useful, offers no account of the latent processes governing action selection, learning, or recovery, leaving clinicians without a principled basis for sequencing interventions when first-line treatments fail.

Recent developments in computational psychiatry offer a framework for this gap. Bayesian models of brain function and the Free-Energy Principle conceptualize perception, action, and learning as hierarchical inference regulated by precision, the confidence assigned to sensory evidence, action policies, motivational priors, and beliefs about environmental volatility.^10,11^ Within this view, psychopathology arises not from isolated deficits but from misallocation of precision across hierarchical levels, producing unstable inference, impaired agency, or maladaptive belief updating.^11,12^

We therefore hypothesized that the heterogeneity of catatonia reflects distinct expressions of failure within a limited set of precision-control domains rather than multiple unrelated mechanisms. We define five domains, each stated in computational terms and then as something a clinician can observe. Sensory precision (πs) is the weight given to incoming sensory evidence: clinically, whether ordinary light, sound and touch arrive as usable information or as noise that provokes withdrawal or agitation. Policy precision (β) is confidence in selecting and executing an action: clinically, whether the patient can carry out an action at all, including one prompted by another person, such as opening the mouth when a spoon is brought. Motivational precision (πm) is the weight on prior preferences that makes an outcome worth pursuing: clinically, whether the patient initiates action without prompting. Fast contextual volatility precision (πv_fast) is the estimate of how quickly the present situation is changing: clinically, whether a minor event is treated as such rather than as a catastrophe. Slow structural volatility precision (πv_slow) is the same estimate over long timescales: clinically, restored sleep-wake structure and a stable account of oneself across contexts and caregivers (Fig. 1).

**Fig. 1.**
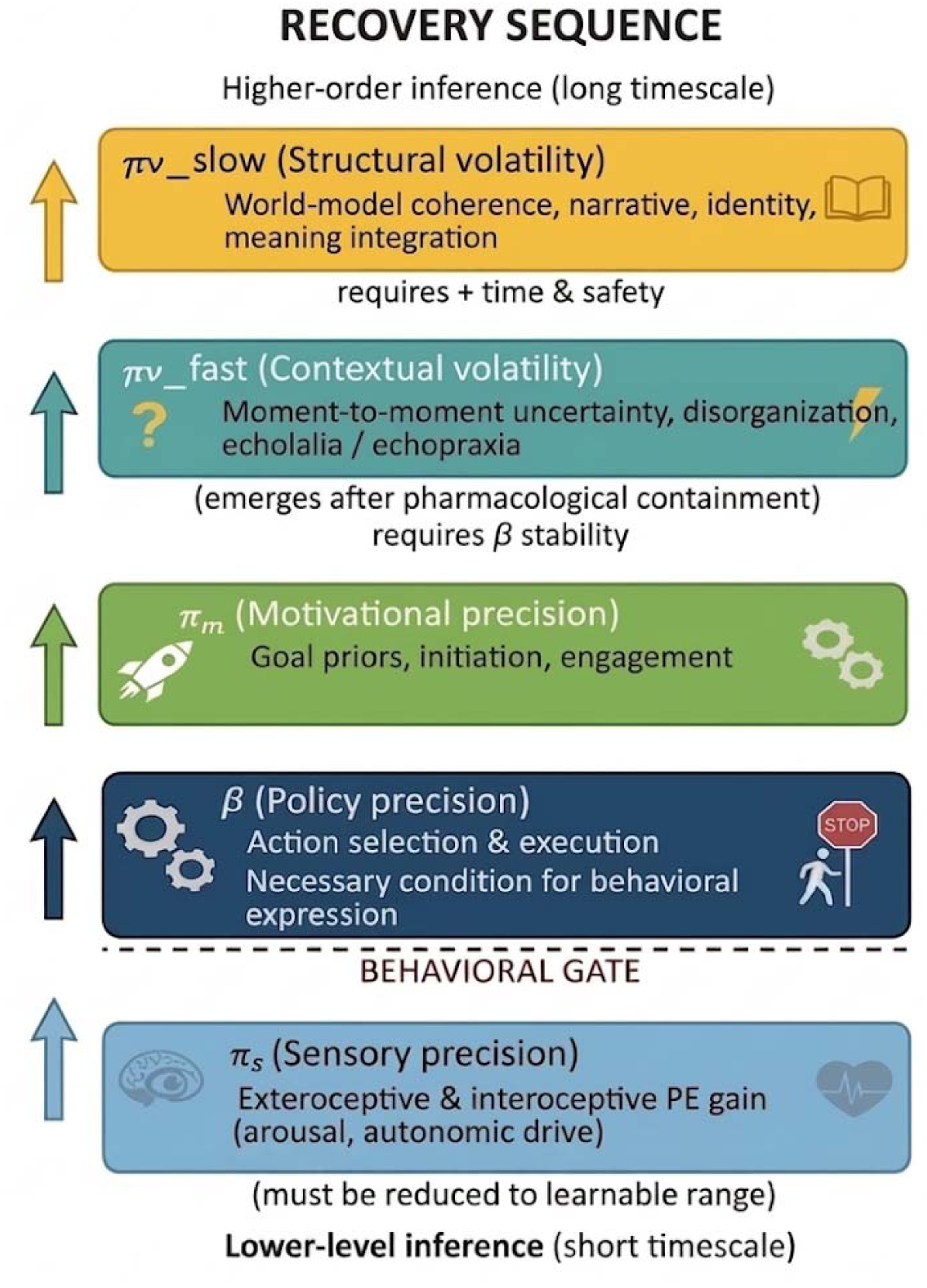
Proposed hierarchical precision model of catatonia. Catatonic phenomena are represented as disturbances of precision weighting across hierarchical levels of inference operating on distinct timescales: sensory precision (πs), gain on exteroceptive and interoceptive prediction errors; policy precision (β), stability of action selection and execution; motivational precision (πm), precision of goal-related priors; fast contextual volatility (πv_fast), moment-to-moment contextual uncertainty; and slow structural volatility (πv_slow), longer-timescale coherence of the generative model. Arrows denote hierarchical dependencies specified a priori from active inference, not linear causality; bracketed annotations mark the prerequisite relationships fixed before milestone coding. The hierarchy is a computational hypothesis intended for prospective testing, not a validated model.

A road analogy may help a clinical reader hold the five together; it describes the different roles and does not by itself establish the recovery order or any treatment effect. Sensory precision is the information coming up from the road surface; policy precision the steering, by which an intended action is carried out; motivational precision the engine, the drive to set off without being pushed; fast contextual volatility precision the satellite navigation (Global Positioning System, GPS), which re-plans the route as the situation changes; and slow structural volatility precision the driver, who keeps the car on track across changing roads and hours. Table 1 maps all DSM-5-TR catatonia criteria onto these domains, with policy precision conceptualized as a cross-cutting gate on behavioural expression.^12,13^

The status of these five quantities determines how the study can be read. The domains are clinically interpretable abstractions informed by active inference and hierarchical precision models. They were not estimated as latent variables from behavioural data; the π and β notation denotes the theoretical construct being indexed, not a fitted parameter.

The order follows from a prerequisite argument: where a lower capacity is absent, the record carries no information about the capacity above it, so the lower capacity must be documented as returning first. This is a prediction about the order in which capacities can be observed to return in a clinical record, conditional on the dependency structure the companion model assumes; it does not by itself show that the underlying function above cannot recover earlier. Of the 120 possible orderings of five domains, those in which a capacity is documented before another capacity that its definition presupposes are excluded by construction; the framework licenses one order of recovery, πs → β → πm → πv_fast → πv_slow. Because the ordering was specified a priori and then used to operationalize the milestones, the analysis that follows demonstrates internal consistency of that scheme with the records; it does not independently establish the hierarchy. The domain definitions, the ordering and the classification of transitions as separable or nested were fixed in writing before any milestone date was assigned, but after the admissions had taken place and with clinical familiarity with the cohort. The study is therefore hypothesis-generating, and the order it reports was formed with knowledge of the clinical course rather than predicted independently of it.

We examined the records of twenty-five consecutive inpatients with catatonia treated during routine care (Supplementary Note 1), and then had every milestone date re-rated by a co-author blind to the hypothesis.

## Methods

### Study design and cohort

This study was conducted as a retrospective, hypothesis-generating clinical investigation based on anonymized medical records. We reviewed the charts of twenty-five inpatients who met DSM-5-TR criteria for catatonia and were treated at Onda-daini Hospital, a psychiatric facility without ECT capability. Baseline demographic and clinical characteristics of the twenty-five patients, including primary diagnosis and catatonia subtype, are summarized in Supplementary Table 1. Five of these twenty-five cases were previously reported in a preliminary case series describing the clinical resolution of catatonia with guanfacine^14^; however, the present study does not extend or reanalyse pharmacological outcomes but instead re-examines all cases within a unified computational framework.

Between April 2021 and March 2026, these twenty-five patients constituted all consecutive catatonia cases managed at the institution, with no exclusions; all reached sustained functional remission and none were transferred elsewhere for ECT. Cases were identified by review of the hospital’s complete inpatient records rather than by diagnostic-code search. Because the hospital has no ECT capability and no external facility accepted transfer for ECT during the study period, treatment-resistant patients were necessarily managed on site to remission rather than transferred out, so the cohort cannot have been depleted of refractory cases through transfer. The corollary is that the cohort contains no non-remitting or relapsing course, and therefore cannot test the framework’s predicted signatures of failed recovery (see Discussion). No experimental intervention, protocolized treatment, or prospective assignment was performed. Clinical management was determined solely by treating physicians according to standard practice. The present analysis represents a secondary interpretation of previously collected clinical data.

### Diagnostic ascertainment and exclusion of medical causes

Catatonia was diagnosed by DSM-5-TR criteria on longitudinal clinical assessment, and psychiatric diagnoses and clinical features, including malignant risk, were established by chart review. Medical causes were investigated in all twenty-five patients by cranial computed tomography, electroencephalography and blood testing, and in no patient did these identify a condition that explained the clinical picture. The hospital has no facility for cerebrospinal fluid analysis or magnetic resonance imaging, and formal neuropsychological assessment and urine drug screening were not performed, so secondary catatonia from an occult cause cannot be excluded with certainty in these patients. The aetiological scope of the framework, and where a secondary cause would be expected to depart from the presentations studied here, are set out in Supplementary Note 2.

### Pharmacological interventions

All treatment was chosen by the treating physicians on clinical grounds during routine care and not by protocol. Every patient received a benzodiazepine (lorazepam in 23, diazepam in one and ethyl loflazepate in one). Pregabalin, for which improvement of catatonia has been reported in a single case,^15^ was added in four patients, and memantine, used adjunctively in refractory catatonia,^7^ in two. Guanfacine, an alpha2A-adrenoceptor agonist, was used in 16 of the 25 patients. Its indication was catatonia itself, after incomplete response to benzodiazepines, including the autonomic instability of malignant catatonia, and not attention-deficit hyperactivity disorder or agitation as such. The rationale was that noradrenergic modulation of prefrontal signal-to-noise might stabilize action selection where sedation alone had not, following a case series from this institution.^14^ The experimental basis is that α2A-adrenoceptor stimulation strengthens prefrontal network activity by inhibiting cyclic AMP–HCN channel signalling.^16^ This rationale is distinct from increasing sedation and does not establish an effect on policy precision, nor efficacy in catatonia. Guanfacine was started at a low dose and titrated in relation to response and blood pressure over a range of 1 to 6 mg daily. An antipsychotic was used during contextual stabilization in every patient, most commonly olanzapine (11 patients), brexpiprazole (5), quetiapine (4), aripiprazole (4), lurasidone (4) or paliperidone (3); valproate was used in three and an antidepressant in four patients. Agents by patient and by milestone are given in Supplementary Table 2, and the change in concurrent agents between baseline and remission is reported in the Discussion.

### Computational framework and inferential strategy

We employed a qualitative, theory-informed analytic framework grounded in Bayesian inference and the Free-Energy Principle. Precision was treated as a latent construct inferred retrospectively from observable clinical signs, behaviours, and functional capacities documented in the medical records, consistent with Bayesian formulations of individual learning under uncertainty.^10,17,18^

No precision parameter was estimated numerically. Three things are therefore kept apart: the theoretical construct (the precision term), its clinical operationalization (the milestone definition, an observable event) and an empirically estimated latent variable, which was not obtained; the five domains are clinical abstractions, not latent variables identified from these data. This concerns what can be estimated from the present records; whether the five parameters are identifiable in principle within a specified model is a separate question, addressed in a companion theoretical work.^19^ DSM-5-TR criteria served as clinical anchors: each was assigned a priori to one or more domains according to the level of inference most directly implicated by the observable sign (Table 1),^12,13^ and the mapping was then used to operationalize the milestones.

### Coding procedure, ambiguity and blinding

Primary coding was performed by one investigator (H.S.), who knew the hypothesized order, and every milestone statistic in the Results that is not labelled as from the blinded re-rating derives from this coding. A second rater, blind to the hypothesis, re-rated all milestone dates after the primary analysis had been completed, under a pre-registered plan (Blinded re-rating of milestone dates, below), and agreement between the two ratings is reported. Assignment required judgement in two places. Some signs are compatible with more than one domain. Agitation, for example, can reflect heightened sensory reactivity, weakened control of action or rapid contextual instability. Such a sign was not assigned to a mechanism: an entry was used only where it met a specific milestone definition, and the alternative reading was recorded. Where the record was ambiguous as to whether a behaviour was prompted or spontaneous, the more conservative assignment was taken, that is, β rather than πm. Worked examples of source entries and their resulting assignments are given in Supplementary Note 3. Slow structural volatility precision (πv_slow) corresponds to no single DSM-5-TR criterion; it was ascertained from documented cross-context stability during remission, as defined below, and not from the criterion set.

### Ascertainment of resolution

Routine records document the presence of a sign more reliably than its resolution. Two rules were therefore applied. A milestone was dated from the first record entry that positively documented the relevant capacity, that is, an entry describing the behaviour itself, rather than from an entry in which the earlier abnormality was merely absent from the note. Where the record did not establish that a domain’s baseline disturbance had been present, the domain was coded as not applicable rather than as resolved; this is why fast contextual volatility precision is missing in three patients of the primary coding. Both rules err towards dating milestones late rather than early. Medical record entries are also more frequent at the acute peak of catatonia than during recovery, so a capacity may have returned some time before the entry that first documents it; neither rule can correct for this.

## Outcome measures

### Primary outcome

Catatonic severity was quantified with the BFCRS^20^. Totals at peak severity and after clinical remission were scored retrospectively from the documented examination findings (23 items, each 0 to 3); items requiring an elicitation manoeuvre that was not documented were scored as absent, so item totals reflect documented findings and may underestimate those signs. Reduction in BFCRS score served as the primary quantitative indicator of catatonia resolution. Because elicited signs requiring a specific bedside manoeuvre may be under-ascertained in routine records, we performed a pre-specified sensitivity analysis on an observational subscale comprising the 16 items scorable without such a manoeuvre (excluding rigidity, waxy flexibility, gegenhalten, mitgehen, ambitendency, grasp reflex, and automatic obedience), repeating the Wilcoxon signed-rank comparison of peak versus remission scores (IBM SPSS Statistics v32). The BFCRS was the only structured instrument in the records; standardized ratings of psychotic or mood symptoms were not part of routine documentation and are not available.

### Secondary functional outcomes

Functional recovery was assessed based on documented restoration of autonomous eating, cooperation with care, mobility, and stabilization of autonomic regulation. Functional remission was defined as near-complete resolution of catatonic features accompanied by restoration of goal-directed behaviour, without escalation of antipsychotic load or use of ECT.

### Sequence analysis of recovery milestones

Five recovery milestones, one per precision domain, were each defined as the first documented, sustained resolution of a domain-specific disturbance present at baseline, rather than as a fixed behaviour, so that heterogeneous presentations could instantiate the same milestone through different observable events. Reduction of sensory precision disturbance (πs) was the first sustained attenuation of sensory-driven reactivity documented at baseline, including autonomic instability, unprovoked agitation, fear-driven withdrawal, or sensory-driven defensive behaviour. Stabilization of policy precision (β) was the first sustained tolerance and execution of structured care, feeding or ambulation without behavioural collapse under external guidance. Restoration of motivational precision (πm) was the first sustained recovery of goal-directed drive, evidenced either by spontaneous, unprompted initiation or by self-sustained continuation and generalization of goal-directed behaviour beyond a prompt without step-by-step re-prompting; engagement requiring a fresh external prompt for each step was classified as β rather than πm. Containment of fast contextual volatility (πv_fast) was the first sustained interval, within a day or across repeated interactions within a day, free of short-timescale (within-interaction, second-to-minute) instability documented at baseline, including echo phenomena, context-inappropriate behaviour, abrupt behavioural branching, or non-psychotic fluctuation in responsiveness and communication, irrespective of the presence of psychotic symptoms or antipsychotic treatment. The milestone indexes the contextual coherence of speech and conduct, not the presence or absence of psychotic content: a patient who retains delusional beliefs but whose speech and behaviour are coherent across an interaction reaches it, and residual psychotic symptoms were not rated with a standardized instrument (their documented course is reported separately). Where no such short-timescale instability was documented at baseline, the domain was coded as not applicable. Slow structural reintegration (πv_slow) was the first sustained period (≥24–72 h) of cross-context stability, defined as a consistent level of functioning maintained across day and night and across at least two distinct contexts or caregivers, including restored sleep–wake structure and non-disrupted eating, conversation, mobility and interpersonal responsiveness.

For each patient, a domain was coded only if its baseline disturbance was documented; otherwise it was recorded as not applicable. The day (counted from the onset of catatonic symptoms, the day of onset being day 1) of each applicable milestone was extracted from the medical record, with the supporting record entry retained for audit; every applicable milestone was reached before discharge in every patient, so no censoring was required. Each milestone was scored in the same way, as a single calendar day identified retrospectively from the chart record. Sustained resolution was judged from subsequent documentation rather than from a uniform assessment schedule, so the stored date is the entry at which the capacity was first positively documented; the interval to its confirmation was not recorded separately, and observation density can therefore affect the dates. This applies to πv_fast as it does to πs, β and πm: each was dated to the first day on which its definition was met, and no domain required the rest of the admission to be observed before it could be dated, so all four were dated on the same footing. Domain milestone dates were coded independently of one another, and the hypothesized order (πs → β → πm → πv_fast → πv_slow) was specified a priori. A companion theoretical paper derives this order formally: where a prerequisite is absent, the likelihood is flat in the dependent parameter, so the prerequisite relations form a chain whose topological ordering is unique and identical to the order tested here. That derivation takes only the five domains as input and does not use the milestone timings reported in this paper.^19^ For each adjacent domain pair, we computed the proportion of patients in whom both milestones were applicable and the observed order matched the prediction; same-day milestones were treated as ties and the rate of same-day co-occurrence was reported. As a global index, we computed Kendall’s coefficient of concordance (W) across patients, with its χ² statistic testing the observed ordering against chance. Ties were resolved by mean ranks and W was corrected for ties. Because same-day co-occurrence is predicted by the ordering hypothesis rather than being a nuisance, deflating the coefficient for ties would penalize the hypothesis for a prediction it makes; the tie-corrected value is therefore reported as the primary index, with the uncorrected value (W = 0.82, χ² = 72.2) given for comparison. We pre-specified that adjacent-pair concordance not exceeding chance would be interpreted as failure to support the hypothesized sequence. All statistical analyses were performed using IBM SPSS Statistics version 32.0 (IBM Corp., Armonk, NY, USA); the descriptive statistics, the coefficient of concordance and Figs. 2 and 3 were independently reproduced in Python with the archived analysis script (see Code availability).

**Fig. 2.**
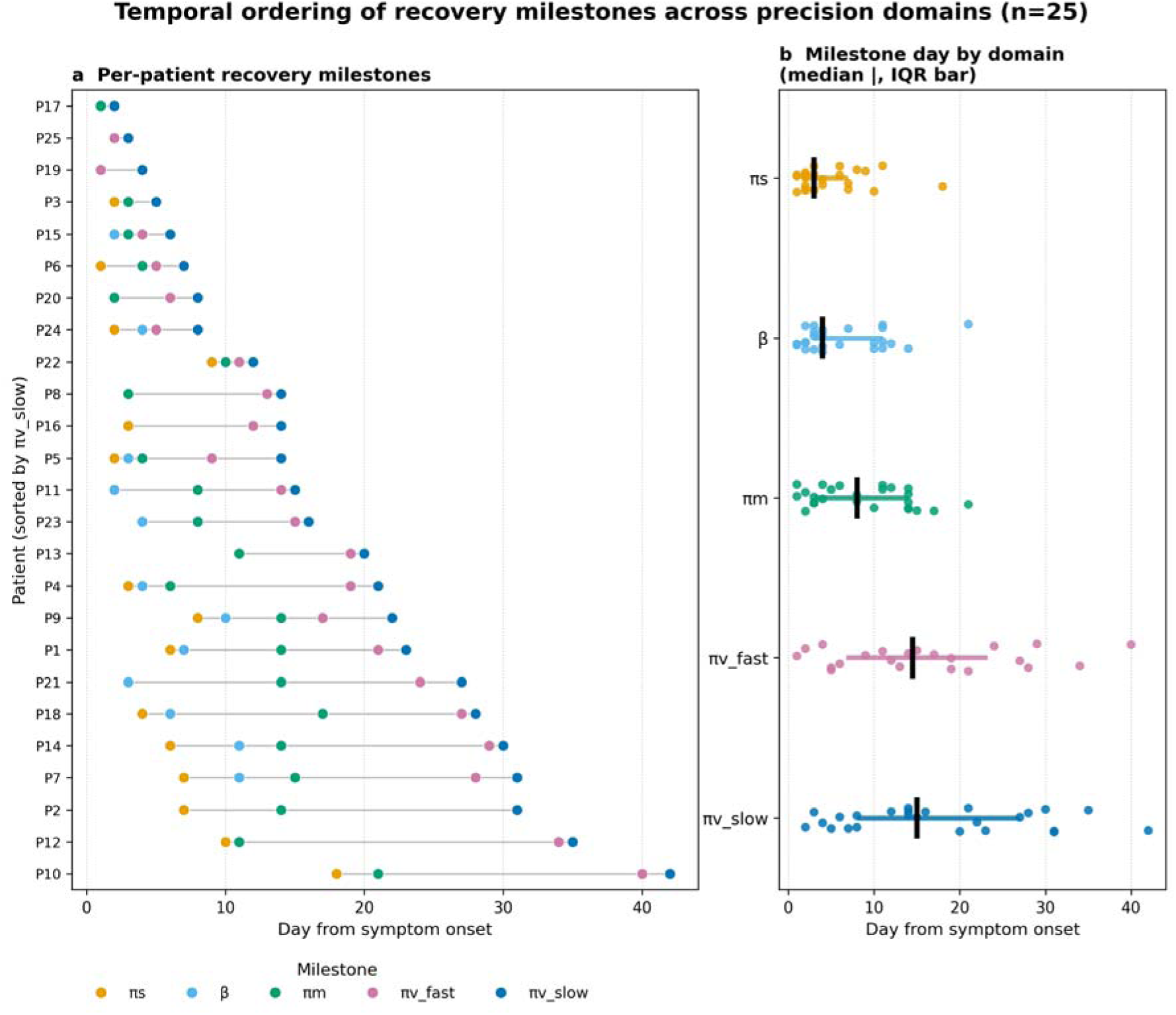
Temporal ordering of recovery milestones across precision domains (n = 25). a, Per-patient milestone days from symptom onset (day of onset = day 1), sorted by πv_slow; each marker is the day a domain’s milestone was reached (πv_fast not applicable in three patients). b, Distribution of milestone day by domain (vertical mark, median; horizontal bar, interquartile range). The predicted order πs → β → πm → πv_fast → πv_slow held without exception in the 15, 12, 18 and 22 patients whose successive milestones fell on different days and therefore carry order information for the four adjacent pairs, and was not violated in any patient when same-day milestones are counted as consistent; early milestones frequently coincided, whereas later transitions were temporally distinct. The four counts refer to the successive adjacent pairs: 15 patients for πs → β, 12 for β → πm, 18 for πm → πv_fast and 22 for πv_fast → πv_slow. The remaining patients in each pair reached both milestones of that pair on the same day and are therefore uninformative about its order rather than discordant with it. Not every patient contributed every milestone: πv_fast was not applicable in three patients, so the two pairs involving it are based on 22 patients. No inversion occurred in any patient at any pair.

**Fig. 3.**
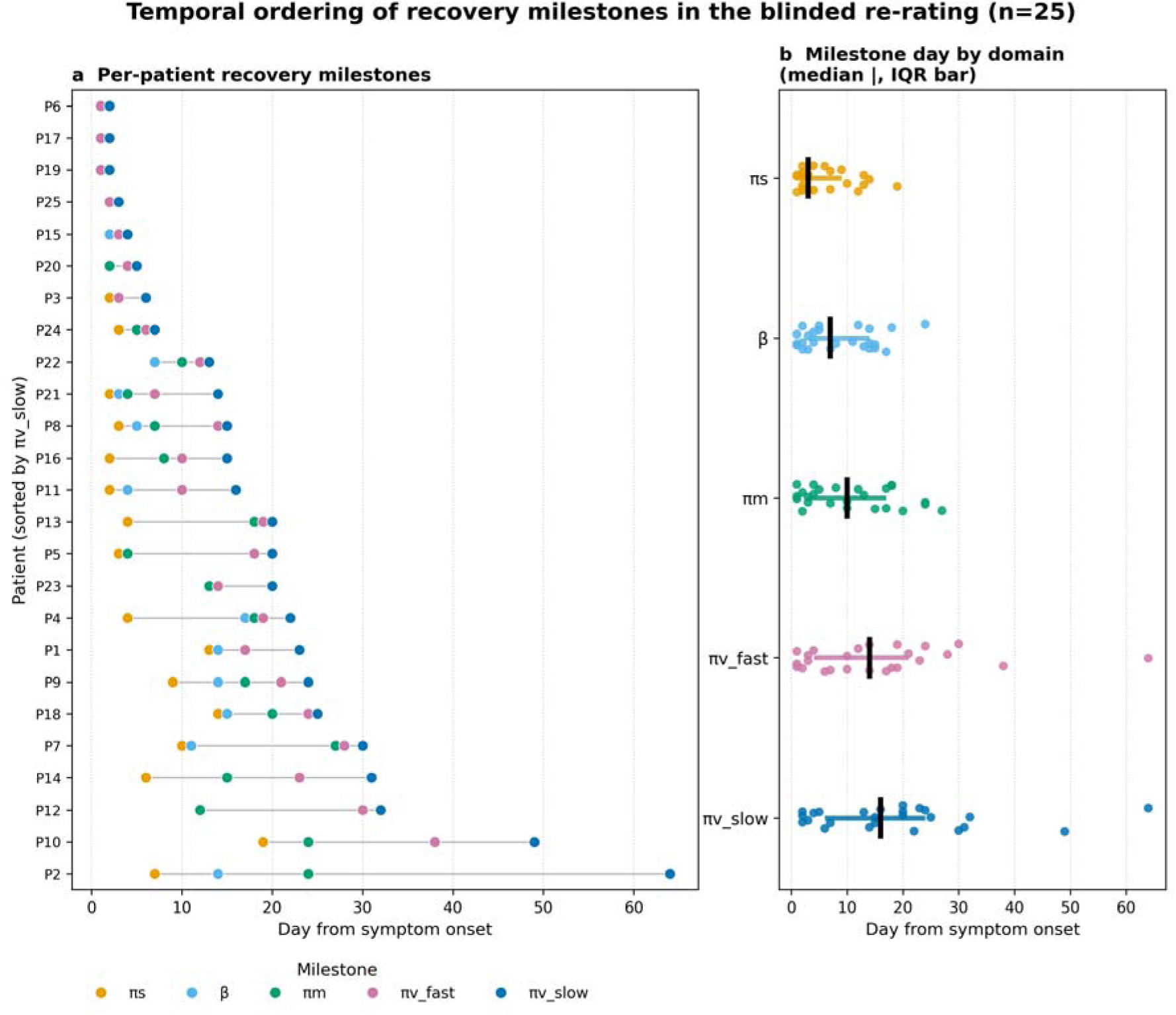
Temporal ordering of recovery milestones in the blinded re-rating (n = 25). Milestone days as rated by the hypothesis-blind co-author (I.M.) from the medical records under the pre-registered plan, shown in the same format as Fig. 2. a, Per-patient milestone days from symptom onset (day of onset = day 1), sorted by πv_slow; each marker is the day a domain’s milestone was reached (every domain was dated in every patient). b, Distribution of milestone day by domain (vertical mark, median; horizontal bar, interquartile range). The predicted order πs → β → πm → πv_fast → πv_slow held without inversion in the 16, 11, 17 and 24 patients whose successive milestones fell on different days for the four adjacent pairs; the remaining patients in each pair reached both milestones on the same day. Agreement with the primary coding is given in Supplementary Table 7 and both raters’ days in Supplementary Table 8.

Adjacent-pair separability was classified a priori from the milestone definitions: a pair was labelled separable if neither definition referenced the capacity indexed by the other (πs → β; πm → πv_fast), and nested otherwise (β → πm, in which motivational precision presupposes executable action; πv_fast → πv_slow, in which the slow-structural window presupposes short-timescale stability). This classification was fixed before ordering analysis. For separable pairs, the observed order is not entailed by the definitions, though this does not establish that the domains are mechanistically independent; for nested pairs, we additionally report inter-milestone latency, which the definitions do not constrain.

### Blinded re-rating of milestone dates

After the primary analysis had been completed, the five milestone dates were re-rated for all 25 patients from the medical records by a psychiatrist at the same hospital (I.M.) who had previously taken no part in the study, had not seen the manuscript, the preprint or the companion theoretical papers, and was not told the hypothesized order or the first rater’s dates. The rater received the five milestone definitions given above, labelled A to E in an order unrelated to the hypothesized sequence, and a rating manual, and entered for each label the first day on which the definition was met, or a code (NA or ND) where it could not be dated. The manual’s rules for doubtful cases err towards dating milestones late, as in the primary coding, and the sheet was locked on return (procedure, codes and label correspondence in Supplementary Note 4). The rater returned each milestone as a day count from the onset date recorded for the primary analysis (day of onset = day 1), so the returned sheet carries no calendar information.

The analysis plan, the rating manual and the label correspondence were deposited at Zenodo^21^ (doi:10.5281/zenodo.23076104) on 1 October 2026, before any rating had been returned, and the deposit was disclosed to M.T. on the same day; it was held under embargo while the ratings were collected and was made public on 16 October 2026. The plan fixed the primary criterion as the number of inversions of the two separable transitions (πs → β; πm → πv_fast) in the blind ratings: a single inversion refutes the pre-specified order, and no threshold for partial agreement was set. The nested pairs (β → πm; πv_fast → πv_slow) are reported descriptively. Agreement between the two raters is summarized by the intraclass correlation coefficient ICC(A,1) with its 95% confidence interval and Bland–Altman limits of agreement for the milestone day, by Cohen’s κ for whether a milestone could be dated at all, and by Kendall’s τ-b for the order of the five milestones within each patient; Kendall’s W computed on the blind ratings is reported beside the primary value as a reproducibility index, without a threshold. Analyses were run in IBM SPSS Statistics v32 and reproduced with a script archived with the figure code (Code availability). The rater was unblinded, and joined the author list, only after the ratings had been locked.

### Course of the underlying disorder

As an exploratory check on whether the milestone order tracks the resolution of the underlying disorder, H.S. extracted from the same records, for each patient, whether the symptoms of that disorder were still documented at discharge. For the 22 patients with schizophrenia or schizoaffective disorder these symptoms were hallucinations and delusions; for the three with a mood disorder they were mood symptoms, with any psychotic features. Where the symptoms resolved, the last entry documenting them was placed relative to the five milestone dates. This extraction was made after the primary analysis, by the unblinded investigator, without a standardized instrument, and is reported descriptively.

### Statistics

All tests were two-sided, with α = 0.05. Peak and remission BFCRS scores were compared within patients using the Wilcoxon signed-rank test; because the scores are bounded ordinal sums containing ties, p values are reported from the tie-corrected normal approximation, and the effect size is r = |z|/√n. Descriptive statistics are given as mean ± standard deviation with median and interquartile range. The χ² statistic accompanying Kendall’s W was evaluated on k − 1 = 4 degrees of freedom in the 22 patients in whom all five milestones were applicable. For each adjacent pair, the direction of the observed order was tested with an exact two-sided sign test restricted to the patients whose two milestones fell on different days and therefore carry order information. No covariates were tested and no adjustment for multiple comparisons was applied: the ordering was specified a priori as a single directional prediction, and the concordance and sign-test statistics are reported as summary indices of that one prediction rather than as independent hypotheses. No Bayesian analysis was performed and no hierarchical or multilevel model was fitted; the design is a within-patient ordinal comparison in which each patient serves as their own reference. Statistics for the blinded re-rating follow the pre-registered plan and are specified under Blinded re-rating of milestone dates.

## Ethics oversight

Ethical approval for this retrospective study was obtained from the Institutional Review Board of Onda-daini Hospital (approval number: onR-10). All procedures were conducted in accordance with the ethical standards of the Declaration of Helsinki. All data were anonymized prior to analysis. Written informed consent for the use of anonymized clinical information and for publication was obtained from patients or their legal representatives in accordance with institutional standards.

## Results

### Cohort characteristics

The 25 patients comprised 13 men and 12 women, aged 55.9 ± 13.9 years (median 57, IQR 46–63, range 26–81; individual ages are given by decade in Supplementary Table 1, to limit re-identification). Primary diagnoses were schizophrenia in 18, schizoaffective disorder in four, bipolar disorder in two and major depressive disorder in one. Catatonia was excited in 11 patients, retarded in 10 and malignant in four. Peak Bush–Francis Catatonia Rating Scale (BFCRS) total was 26.3 ± 5.6 (median 26, interquartile range (IQR) 23–30, range 14–34). Per-patient characteristics are given in Supplementary Table 1 and per-patient BFCRS totals in Supplementary Table 3, and the prevalence of each BFCRS item at peak in Supplementary Table 4.

### Treatment context

Initial management documented in the records was empirically selected and guideline-concordant, and reduction of visible agitation or sedation alone did not reliably coincide with functional reintegration: when aroused, many patients remained incoherent, excited, manneristic or frozen, and BFCRS totals frequently stayed elevated. Agents of distinct classes were in use around the milestones (Supplementary Table 2). The same milestone order was documented irrespective of which agents were in use, and narrative coherence and cross-context consistency recovered last.

### Summary of outcomes

Across all twenty-five cases, mean BFCRS score decreased from 26.3 ± 5.6 at peak to 2.0 ± 2.4 (median 0, interquartile range (IQR) 0–4) after remission (Wilcoxon signed-rank test, z = 4.38, p = 1.2 × 10; Supplementary Table 3); all improved, with 13 reaching 0 and 17 scoring 3 or below, and all achieved sustained functional remission. A sensitivity analysis confined to observationally scorable signs left this reduction essentially unchanged (Methods; note to Supplementary Table 3), so it was not an artefact of untested elicited items. Benzodiazepine doses were reduced in many cases after remission. This convergence across heterogeneous presentations supports a shared temporal structure of recovery from catatonia.

### Ordered recovery across precision domains

The predicted order (πs → β → πm → πv_fast → πv_slow) held without a single inversion in every patient carrying order information for a given pair, and, when same-day milestones are counted as consistent, in every patient in whom both milestones of a pair were applicable. Because two adjacent transitions are partly definitional (see below), the primary test rests on the two operationally separable transitions (πs → β; πm → πv_fast), and the global concordance across all five milestones (tie-corrected Kendall’s W = 0.91, uncorrected 0.82; χ²(4) = 80.4, p = 1.5 × 10 ¹; n = 22) is reported as a summary index rather than the principal evidence. Median time from symptom onset increased monotonically across domains: πs, 3 days (IQR 2–7); β, 4 (3–11); πm, 8 (3–14); πv_fast, 14.5 (6.8–23.2); πv_slow, 15 (8–27) (Fig. 2; Supplementary Table 5). Pairwise, the number of patients carrying order information (no same-day tie) was 15 for πs → β, 12 for β → πm, 18 for πm → πv_fast and 22 for πv_fast → πv_slow, and in each of these the direction was the predicted one (exact sign test, p = 6.1 × 10, 4.9 × 10, 7.6 × 10 and 4.8 × 10, respectively); counting same-day ties as consistent, agreement was 25/25, 25/25, 22/22 and 22/22, respectively (Supplementary Table 6). πv_fast was coded not assessable in three patients. All three had received an antipsychotic from the start of treatment, and the baseline record was ambiguous as to whether short-timescale instability had been present.

For the two separable pairs (πs → β; πm → πv_fast), the order is not entailed by the milestone definitions, and both held without inversion. For the two nested pairs (β → πm; πv_fast → πv_slow), whose definitions share a rank-order dependency, the informative quantity is instead the inter-milestone latency, which the definitions do not constrain and which was non-zero in 12/25 and 22/22 patients, respectively.

### Blinded re-rating

In the blinded re-rating (I.M.), neither separable transition was inverted in any patient carrying order information (πs → β, 0 of 16; πm → πv_fast, 0 of 17), so the pre-specified order was not refuted. For the nested pairs, inversions were 0 of 11 for β → πm and 0 of 24 for πv_fast → πv_slow (Fig. 3; Supplementary Table 7). Agreement between the blind and primary milestone days, estimated with ICC (A,1), was 0.71 (95% CI 0.46 to 0.86) for πs, 0.72 (0.42 to 0.87) for β, 0.75 (0.50 to 0.88) for πm, 0.90 (0.76 to 0.96) for πv_fast and 0.84 (0.66 to 0.92) for πv_slow; Bland–Altman mean differences and limits of agreement are given in Supplementary Table 7. On applicability, the blind ratings contained no NA or ND entry, whereas the primary coding had three (all πv_fast); in four domains, therefore, both raters dated every patient. For πv_fast the blind rater dated all 25 patients, including the three coded not assessable in the primary analysis; κ was 0 because, with every patient in one category for the blind rater, the observed agreement (22 of 25) equaled the agreement expected by chance (Supplementary Table 7). The median within-patient Kendall’s τ-b between the two raters’ orders was 0.95 (range 0.67 to 1.00; n = 25). Tie-corrected Kendall’s W in the blind ratings was 0.89 (n = 25) against 0.91 in the primary coding. Both raters’ day for every patient and domain are listed in Supplementary Table 8.

### Relation to the course of the underlying disorder

In five patients (cases 6, 7, 11, 12 and 13; four with schizophrenia and one with schizoaffective disorder), hallucinations and delusions remained documented throughout the admission and at discharge, while all five milestones were attained in the predicted order of recovery from catatonia. In the three patients with a mood disorder (cases 23 to 25), the underlying disorder remitted after πv_fast. In the remaining 17 patients the last documentation of psychotic symptoms fell on the day coded as πv_slow. In no patient did the underlying disorder remit before πv_fast, and in 22 of 25 its symptoms were documented up to the day of πv_slow or beyond. The coincidence between the last documentation of psychotic symptoms and πv_slow in 17 patients was recorded by the same unblinded rater.

## Discussion

We reinterpret catatonia as a structured disturbance of hierarchical precision control underlying its heterogeneous motor and behavioural manifestations and constraining recovery in a state-dependent manner. Across cases, phenomenology and recovery could be parsimoniously organized around five latent precision domains: sensory (πs), policy (β), motivational (πm), and fast and slow volatility precision (πv_fast, πv_slow).

Fig. 1 summarizes this organization. A key feature is the temporal asymmetry between fast contextual and slow structural volatility, which explains why attenuation of acute symptoms frequently failed to produce functional reintegration and why recovery, when it occurred, unfolded in a delayed yet reproducible sequence.

The ordering is a prediction stated so as to fail. Five domains admit 120 possible orderings; the framework licenses one, fixed before the milestones were coded and derived independently of them.^19^ More generally, a dependency graph fixes the admissible recovery orders exactly, as the set of its linear extensions, so that a chain such as this one admits a single order; the result holds for any acyclic dependency structure.^22^ Within the applicability conditions established there, namely the specified model class, recovery law and gating-dominated rate regime, a single patient in whom a separable transition inverts (organized action stabilizing before sensory-driven instability abates, or contextual instability resolving before goal-directed initiation returns) would falsify the proposed ordering; outside those conditions such an inversion is permitted rather than disconfirming. None occurred here. Because the cohort remitted uniformly, however, it could not test the framework’s predictions for failed recovery, which remains the sharper test. Companion theoretical work derives them: the same dependency structure, coupled to evidence-dependent bistability, yields self-sustaining recovery, maintained remission after support is withdrawn, or stable partial recovery and abrupt collapse in one construction.^23^

Retrospective review showed that stabilization of organized action repeatedly coincided with clinical turning points. Experience-based dose adjustments, both increases and reductions, often attenuated motor activity or autonomic arousal while patients remained fragmented, frozen or incoherent. By contrast, stabilized action execution was associated with tolerance of structured activity, rest and care without behavioural collapse. As organized action stabilized (β), motivational initiation re-emerged (πm). To assess whether recovery was driven by escalating antipsychotic exposure, we compared the regimen at remission with baseline: concurrent agents had decreased in 9 patients, were unchanged in 14, and increased in only 2, consistent with recovery accompanying stabilization of organized action (β) without requiring escalation of dopaminergic blockade. Counts of concurrent agents do not measure cumulative exposure, dose, or receptor occupancy,^24^ so this comparison constrains escalation rather than quantifying dopaminergic blockade. Containment of fast contextual volatility (πv_fast) followed, with slow structural reintegration (πv_slow) consolidating last as cross-context coherence stabilized within a sustained behavioural envelope, not pharmacologically accelerated.

Guanfacine has been reported as a treatment option,^14^ and it was in use in most of this cohort around the time organized action stabilized. In the nine patients who were not treated with guanfacine the order held without inversion, so the sequence is not dependent on alpha_2_-adrenoceptor agonism; the claim concerns the gating role of organized action, not pharmacology. Some presentations remit rapidly with benzodiazepine monotherapy, plausibly lower-complexity states in which attenuating excessive sensory precision suffices; the cases here instead required restoration of organized action.

Two existing accounts of catatonia bear directly on the domain assignments used here. The first is motor: several catatonic signs are better explained by disturbed motor control than by sensory gain, and stereotypies in particular are usually attributed to basal ganglia circuits that store overlearned movement patterns, while waxy flexibility has been linked, though not established, to proprioceptive processing.^25,26^ Reduced benzodiazepine-receptor binding in the left sensorimotor cortex in akinetic catatonia, and delayed late movement-related cortical potentials with an abnormal response to lorazepam,^27,28^ locate the disturbance in the motor circuits that the policy and sensory domains are taken to index; these findings are consistent with the assignments used here but do not test them. The second account is longitudinal: symptom change under benzodiazepines and under ECT has been characterized in previous cohorts, including differential response across symptom clusters.^29,30^ Those studies describe how much and how fast catatonic signs improve, whereas the present question is which capacity returns before which. The two lines of research are complementary rather than competing. A direct comparison would require item-level longitudinal ratings at a sampling density that these records do not have.

All 25 patients remitted without ECT, but the cohort cannot show that following this order improves outcome relative to timely ECT, nor that delaying ECT in favour of the order is safe for a patient in whom ECT is indicated. The framework is not an argument for withholding ECT. The comparison of order-guided sequencing against timely escalation requires a prospective design in a setting where ECT is available.

After the primary analysis, a co-author blind to the hypothesis re-rated every milestone date from the medical records according to a pre-registered plan that fixed a single inversion of a separable transition as the refutation criterion. No inversion was found, and agreement on dates was moderate to good, with ICC (A,1) from 0.71 to 0.90 (Results). This shows that the sequence can be recovered from the record by an independent rater, not that it holds outside this cohort; and both raters read the same documentation, so a bias in what was recorded, as distinct from how it was read, is not excluded. Structured psychopathological information can now be extracted from clinical narratives by natural language processing; an independently specified extraction rule applied to the same records would test whether the domain assignments depend on a human coder at all, which has not been done.

The order did not track the resolution of the underlying disorder. In five patients hallucinations and delusions were still documented at discharge after all five milestones had been passed in order, and in none of the 25 did the underlying disorder remit before πv_fast. Standardized ratings of psychotic symptoms were not available, and a scale such as the PANSS^31^ would have been difficult to apply for this purpose: several of its items overlap with the phenomena the BFCRS records, and most presuppose an interview that the acutely catatonic patient cannot give. The record-based course reported here is descriptive and unblinded; what it supports directly is that remission of hallucinations and delusions was not required for any of the five milestones to be attained.

Recovery trajectories were not linear but recurrently reinforcing, formalized in Fig. 4 as the Saito Loop. The term denotes recurrence in the parameters that govern recovery, not a cycle in the recovery trajectory itself: the state does not return to πs, and the sequence is not reiterated once πv_slow is reached. Each domain’s stabilization supplied the evidence the next required, from learnable sensory gain through executable action and sustained initiation to contextual and then structural coherence. Disruption at any point would predict non-response, premature escalation, or relapse. However, although causal inference is precluded by the retrospective design, convergence toward a shared phase structure across treatments with distinct modes of action (GABAergic, noradrenergic, dopaminergic/serotonergic, and glutamatergic) is more consistent with a state-dependent constraint on hierarchical inference than with a drug-specific effect. It is a computational hypothesis grounded in hierarchical active inference,^32^ whereby higher-level structural beliefs, once stabilized, exert recurrent top-down constraints on lower-level precision allocation through ongoing message passing between levels.^33,34^ Neurophysiologically, precision-weighted prediction errors at distinct hierarchical levels are encoded by dissociable midbrain and basal-forebrain substrates during sensory learning,^35^ which is consistent with the general principle of level-specific precision on which the loop rests, though it does not test the loop’s particular allocation. Formally, the recurrence in the loop is threshold-level rather than state-level: accumulated slow-timescale evidence enters only through the threshold at which each lower domain stabilizes, so that the trajectory converges to the recovered configuration rather than to a cycle, and that configuration becomes self-sustaining. The companion theoretical work derives this feedback rather than assuming it.^23^ The ordering also runs along the sensorimotor-to-association axis of cortical maturation, primary sensory and motor regions maturing before association cortex.^36^ We note the correspondence without claiming that either explains the other: the present account fixes recovery order from dependency structure and is silent about anatomical arrangement, so whether the two share a common pathway remains open.

**Fig. 4.**
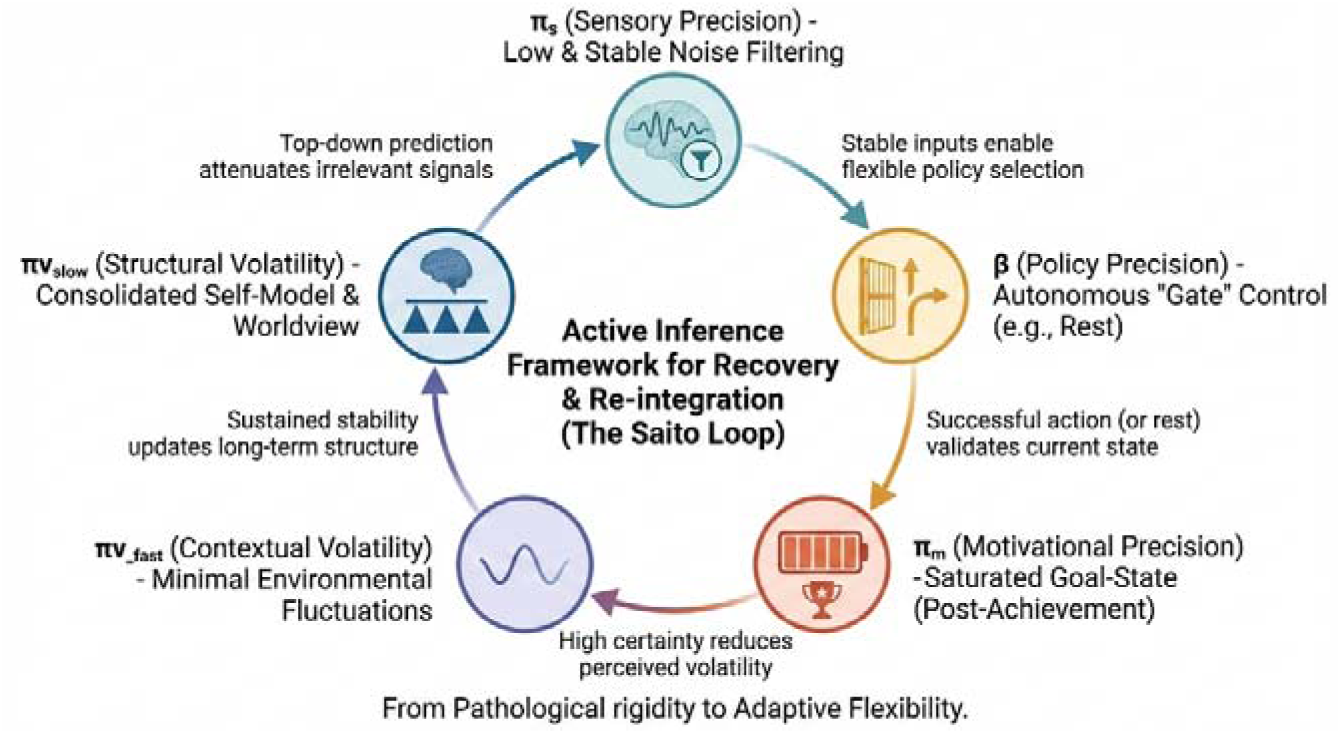
The Saito Loop: recurrent recovery dynamics across hierarchical precision domains. A proposed state-dependent recurrent recovery dynamic, specified a priori from hierarchical active inference, linking the five precision domains across timescales. Recovery trajectories entered the dynamic when excessive sensory prediction-error gain fell into a learnable range (πs), consistent with active inference accounts of sensory attenuation. Stabilization of policy precision (β) was associated with coherent action selection, adaptive rest and predictable engagement with care; sustained execution or rest supplied repeated low-noise evidence from which spontaneous goal-directed initiation re-emerged (πm), and sustained initiation supplied the within-context sampling from which perceived contextual instability fell (πv_fast). Over longer timescales, consolidation of higher-level structure (πv_slow) exerted increasing top-down constraint on lower-level precision allocation. The loop is a computational hypothesis, not a treatment sequence or a literal temporal cycle. The arrow from πv_slow to πs denotes a threshold-level constraint on lower-level precision allocation, not a return of the state to its starting point; once πv_slow is reached the sequence is not reiterated.

Fast contextual volatility appeared as moment-to-moment disorganization, echo phenomena, and context-inappropriate branching over short timescales, whereas slow structural volatility appeared as narrative coherence and cross-context consistency, recovering last. The separation between the two was small in this cohort (median 14.5 versus 15 days, with overlapping interquartile ranges), so these data support the ordering rather than a large temporal gap; stronger claims await prospective data. Slow structural reintegration emerged only after sustained containment of fast volatility (Fig. 1).

Integrating these observations yields a unified model, synthesized in Fig. 5 as “Computational titration”: the state-contingent alignment of hierarchical precision domains across a single traversal of the recurrent loop (Fig. 4).

**Fig. 5.**
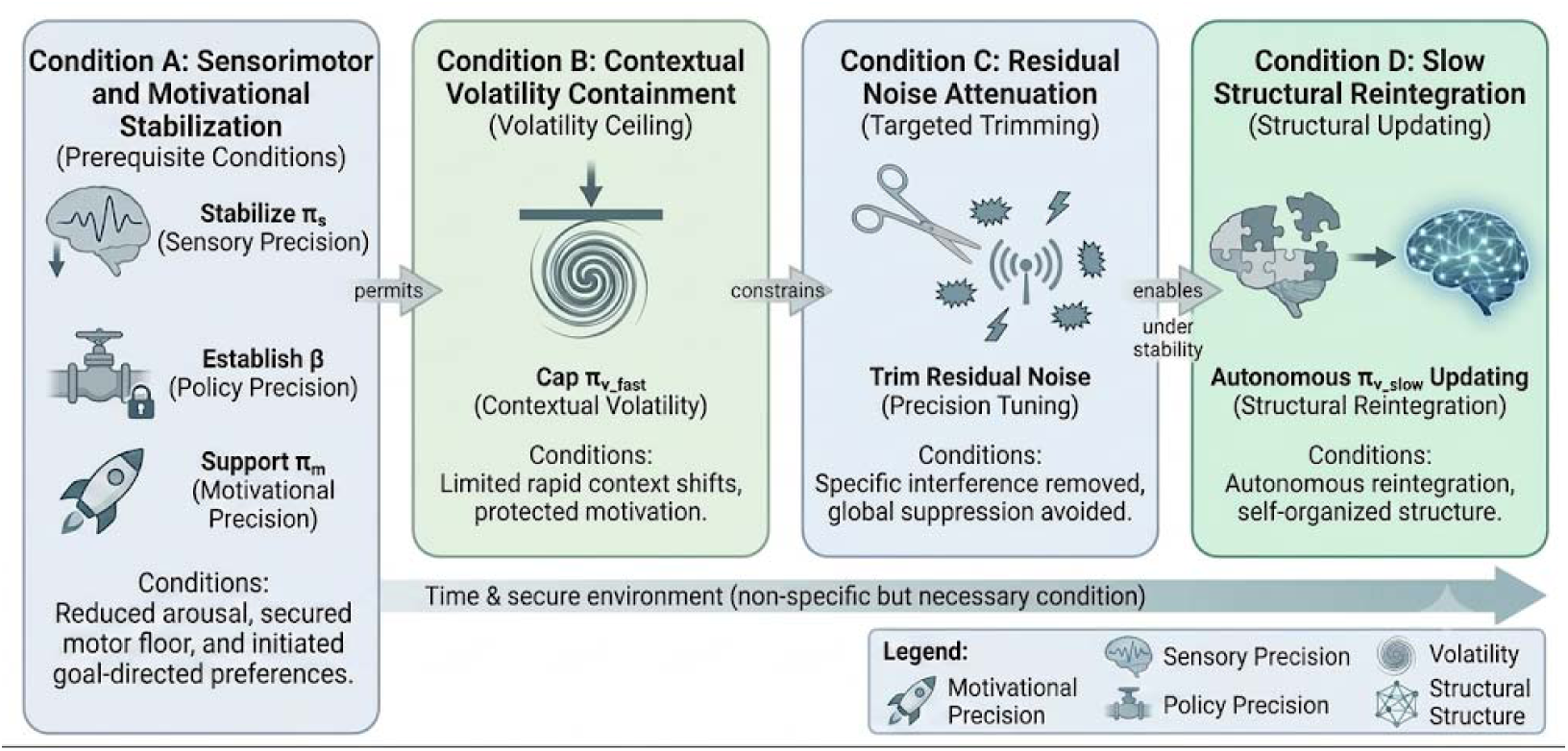
Computational titration: state-contingent precision alignment in catatonia. The constraint structure underlying recovery, specified a priori from the dependency structure of hierarchical active inference, against which the trajectories of the twenty-five cases were examined. Arrows denote conditional dependencies fixed before milestone coding, indicating the state-contingent structure within which stabilization of one precision domain was associated with the permissibility or facilitation of recovery in others; they are not causal, prescriptive or protocolized relationships, and the diagram is not a treatment algorithm, decision tree or clinical recommendation. Domains as in Fig. 1.

Symptom severity alone did not capture the temporal structure of recovery. The ability to taper benzodiazepines after remission is interpretable as a self-sustaining reconfiguration of hierarchical precision control rather than continued dependence on global sensory suppression. It offers a model for apparent treatment failures, paradoxical responses and delayed recoveries. The framework further generates a directly testable prediction for prospective study: once a stable recovery dynamic is established, further pharmacological escalation may be unnecessary or even counterproductive, and controlled reduction of residual agents once organized action has stabilized may itself facilitate recovery.

Whether the framework is general or specific to psychosis-associated catatonia turns on its applicability conditions, which do not reference aetiology. Medical, epileptic, hypoxic, and autoimmune causes account for a substantial share of hospital catatonia, and first-episode psychosis cohorts have been screened for the neuronal autoantibodies that underlie the autoimmune form.^37,38^ Its two dynamical conditions, a common impaired onset and a deficit-closing drive, are where secondary catatonia should depart: a focal or receptor-selective insult need not impair the five domains uniformly, and an active insult opposes closure of the deficit. Non-uniform onset weakens rather than abolishes the prediction, permitting a restricted set of inversions.^19,22^ The ordering should hold after the cause is removed, not during its removal. Supplementary Note 2 develops this, where the single major depressive case falls, and how memantine differs from NMDA receptor autoantibodies.

These findings should be read against three limitations. First, the analysis is retrospective, single-centre, based on a small (n = 25) and uniformly remitting cohort (precluding any test of the framework’s failure predictions). The primary milestone coding was drawn from records not generated for this purpose and performed by a single investigator with knowledge of the hypothesis, and the course of the underlying disorder was extracted by the same investigator without a standardized instrument. The blinded re-rating addresses rater bias but not the dependence of both ratings on the same records. Causal inference is precluded, and replication in independent cohorts will be required. The cohort reflects what a psychiatric hospital without ECT admits, so generalization to medical, epileptic and autoimmune catatonia is untested. Second, the framework is partly confirmatory by construction: because the ordering was specified first and then used to define the milestones, the nested pairs can only show internal consistency, and the test of the order rests on the two separable pairs; early milestones often fell on the same day, which leaves fewer informative observations, although strict temporal separation was nonetheless observed in a substantial proportion of patients. Third, the order in which capacities are first documented need not be the order in which they recover. A capacity cannot be seen until the capacity below it has returned (goal-directed behaviour, for example, can only be identified once organized action is observable), so it may have recovered earlier without leaving a trace in the record; likewise, a capacity that is easier to document may appear to return earlier without having done so. Separating true constraints from these observational dependencies will require prospective, pre-registered validation with predefined criteria, blinded raters, and independent multi-site cohorts. Ordered timing also does not by itself establish mechanistic independence: separable milestones may co-vary through a shared clinical driver (for example, reduced reactivity permitting tolerance of care), so common-cause confounding cannot be excluded.

## Conclusion

Catatonia may be interpreted as a disorder of hierarchical precision control in which disruption of organized action is a key constraint during non-recovery. Recovery coincided not with symptom suppression but with stabilization of organized action preceding motivational initiation, followed by contextual and structural reintegration, across heterogeneous presentations and different treatments. The framework defines testable constraints on sequenced recovery, best understood not as a linear cascade but as a recurrent, self-stabilizing loop (Fig. 4); its principal contribution is a falsifiable recovery ordering, carried primarily by the two operationally separable transitions, that prospective, blinded studies can test. The order held again when every milestone was re-rated by a co-author blind to the hypothesis. Prospective studies are needed to test this order of recovery from catatonia, with blinded ratings in settings where ECT is available.

## Acknowledgements

We appreciate the contributions of all patients and their families to this study. The work was conducted in accordance with the ethical standards of the Declaration of Helsinki and in accordance with the Japanese guidelines on research ethics.

## Funding

No funding was received for this work.

## Author contributions

H.S. conceptualized the study, performed the primary coding and wrote the first draft of the manuscript. I.M. performed the blinded re-rating. Y.T., A.T., J.T., R.A. and M.T. contributed to the modification of the manuscript. All authors contributed to and have approved the final manuscript.

## Competing interests

The authors declare no competing interests.

## Data availability

The data supporting the findings of this study consist of anonymized clinical observations and symptom ratings derived from routine clinical care, including longitudinal BFCRS assessments.

These data are not publicly available due to their clinical nature and the potential risk of patient re-identification but are available from the corresponding author upon reasonable request and subject to institutional and ethical approval. The day counts from onset of both raters, which carry no calendar information, are given for every patient in Supplementary Table 8 and in the public repository named under Code availability; the medical records behind both ratings are not public. The pre-registered analysis plan, the rating manual given to the blind rater and the label correspondence are publicly available at Zenodo^21^ (doi:10.5281/zenodo.23076104); the analysis script is archived with the figure code (Code availability).

## Code availability

The analysis and figure code (catatonia_figure2.py) is openly available at https://github.com/entrance4-png/catatonia-precision-order under the MIT licence and is archived under a persistent identifier at Zenodo.^39^ From the two milestone sheets, which are included in the repository, it reproduces every descriptive statistic, concordance value and panel of Figs. 2 and 3 reported here, and prints both the tie-corrected and the uncorrected coefficient of concordance; these values were computed in IBM SPSS Statistics v32 (Methods) and reproduced with this script. The script for the blinded re-rating (blind_rerating_analysis.py), which reproduces the inversion counts, sign tests, Kendall’s W, ICC, Bland–Altman and κ statistics from the two rating sheets, is archived in the same Zenodo deposit.

## Tables

**Table 1.** Mapping DSM-5-TR catatonia criteria to precision domains.

| DSM-5-TR criterion | Proposed computational interpretation | $\beta$ gate | Clinical readout | Reference |
| --- | --- | --- | --- | --- |
| Stupor | $\beta\downarrow$ and/or $\pi m\downarrow$ | Direct | Marked reduction of spontaneous or reactive movement | Friston 2012<sup>12</sup> |
| Catalepsy | $\pi s\uparrow$ (motor + interoceptive), $\beta$ instability | Often | Posture passively induced and maintained against gravity | Brown 2013<sup>40</sup> |
| Waxy flexibility | $\pi s\uparrow$ (proprioceptive) | Often | Slight, even resistance to repositioning | Brown 2013<sup>40</sup> |
| Mutism | $\pi m\downarrow \pm \beta\downarrow$ | Often | Minimal or absent verbal output | Adams 2013<sup>41</sup> |
| Negativism | $\beta\downarrow + \pi m\downarrow$ | Direct | Resistance or lack of response to instruction | Adams 2013<sup>41</sup> |
| Posturing | $\pi s\uparrow$ (motor set) | Indirect | Active maintenance of abnormal posture | Brown 2013<sup>40</sup> |
| Mannerism | $\pi v\_fast\uparrow$, unstable $\beta$ | Direct | Bizarre or caricatured actions | Schwartenbeck 2015<sup>13</sup> |
| Stereotypy | $\beta$ narrowing + $\pi v\_fast\uparrow$ | Direct | Repetitive, non-goal-directed movements | Friston 2012<sup>12</sup> |
| Agitation | $\pi s\uparrow + \beta\downarrow$ and/or $\pi v\_fast\uparrow$ | Direct | Excessive motor activity without external trigger | Adams 2013<sup>41</sup> |
| Grimacing | $\pi s\uparrow$ (interoceptive/somatic) | Indirect | Sustained facial contortions | Brown 2013<sup>40</sup> |
| Echolalia | $\pi v\_fast\uparrow + \beta\downarrow$ | Direct | Imitative speech | Schwartenbeck 2015<sup>13</sup> |
| Echopraxia | $\pi v\_fast\uparrow + \beta\downarrow$ | Direct | Imitative movements | Schwartenbeck 2015<sup>13</sup> |
| Autonomic instability (not a DSM-5-TR criterion) | $\pi s\uparrow$ (interoceptive) | Indirect | Sweating, tachycardia, vital-sign variability | Adams 2013<sup>41</sup> |

$\pi s$ , sensory precision; $\beta$ , policy precision; $\pi m$ , motivational precision; $\pi v\_fast$ , fast volatility precision; $\pi v\_slow$ , slow volatility precision; $\uparrow/\downarrow$ , increased/decreased precision. $\beta$ gate: Direct, $\beta$ primarily implicated; Indirect, $\beta$ secondarily involved; Often, variably involved. Autonomic instability is not one of the 12 DSM-5-TR criteria and is listed as an additional clinical sign. The final column gives the source of the general precision-theoretic reading applied to each class of sign; no cited study tested the assignment of that sign, and the assignments are the authors' interpretation.

All 12 DSM-5-TR catatonia criteria, and autonomic instability as an additional clinical sign, are mapped onto five computational precision domains derived from Bayesian inference and the Free-Energy Principle: sensory precision (πs), policy precision (β), motivational precision (πm), fast/contextual volatility (πv_fast), and slow/structural volatility (πv_slow). These mappings are grounded in established active inference formulations that relate specific clinical phenomena to alterations in precision weighting across sensory, motor, and policy hierarchies, rather than being derived from post hoc symptom–drug associations.

Policy precision (β) is highlighted as a cross-cutting gate on behavioural expression. Across criteria, sustained catatonic expression is hypothesized to require collapse or instability of β, which governs confidence in selecting and executing action policies. Abnormalities in πs or πv may be sufficient to generate instability or aberrant behaviour, but without β collapse such disturbances remain transient and behaviourally recoverable.

The mapping of DSM-5-TR catatonia criteria to precision domains was based on the clinical level at which each sign becomes observable within a hierarchical active inference architecture, drawing on established models rather than introducing de novo constructs.^10,11,17^ In this framework, DSM criteria were not treated as independent symptoms with one-to-one mechanisms, but as behavioural readouts of disturbances in precision weighting across sensory, policy, motivational and volatility-related levels of inference. Signs dominated by autonomic arousal, grimacing, defensive withdrawal or unprovoked agitation were assigned primarily to sensory precision (πs), because they indicate excessive gain on exteroceptive or interoceptive prediction errors. Signs involving failure to initiate, sustain or flexibly select actions, including stupor, negativism and stereotyped behavioural narrowing, were assigned to policy precision (β), reflecting reduced confidence in action policies and impaired psychomotor control. Mutism and loss of spontaneous goal-directed engagement were mapped to motivational precision (πm), as these signs reflect impaired precision over prior preferences sufficient to initiate action. Echo phenomena, mannerisms and context-inappropriate behaviours were mapped to fast contextual volatility (πv_fast), because they indicate unstable moment-to-moment contextual inference and rapid behavioural switching. Slow structural volatility (πv_slow) was not mapped to a single DSM criterion, but to the delayed recovery of cross-context coherence, narrative continuity and stable interpersonal engagement during remission. Thus, DSM-5-TR criteria served as observable anchors for retrospective coding, whereas precision domains provided the latent computational structure used to interpret recovery trajectories. Several criteria load onto more than one domain, and these assignments are interpretive rather than unique: agitation, for example, is attributed jointly to elevated sensory precision, reduced policy precision, and elevated fast contextual volatility (πs↑ + β↓ and/or πv_fast↑), and manneristic or context-inappropriate behaviours to elevated fast volatility interacting with unstable policy selection, so plausible alternative assignments exist for such multi-determined signs. The ordering hypothesis is nonetheless robust to this ambiguity, because it is defined over the recovery of each domain rather than over the domain attribution of any single criterion: reassigning an individual sign changes which milestone a given behaviour helps index, not the predicted sequence of domain stabilization. Criteria whose attribution is least ambiguous (e.g., autonomic instability → πs; stupor → β/πm) anchor the milestones most heavily, whereas multi-determined signs contribute corroboratively rather than decisively.

## Notes

### Competing Interest Statement

The authors have declared no competing interest.

### Clinical Protocols

https://doi.org/10.5281/zenodo.23076104

### Summary of Updates

Version 2 adds a pre-registered blinded re-rating of all milestone dates. A psychiatrist at the same hospital who had taken no part in the study and was not told the hypothesized order (Issei Murata, now a co-author) re-rated every milestone for all 25 patients from the medical records under random labels; the analysis plan, rating manual and label correspondence were deposited at Zenodo (doi 10.5281/zenodo.23076104) on 1 October 2026 before any rating was returned. The blind ratings contained no inversion of either separable transition. Results are shown in a new Figure 3 and Supplementary Tables 7 and 8, with the procedure in Supplementary Note 4. A new section reports the course of the underlying disorder relative to the milestones. The Introduction now states each domain in clinical terms, and the Methods describe the coding procedure, ascertainment of resolution and the five milestone definitions in more detail. The Discussion adds paragraphs on existing accounts of catatonia, the relation to ECT and reproducibility of the coding. The study is described throughout as hypothesis-generating. Author list, affiliations, Supplementary files and analysis code updated.

