## Supplementary Information (Tables 1-8, Notes 1-4) for "Recovery from catatonia follows a hierarchical precision order"

**Supplementary Table 1.** Baseline characteristics and catatonia subtypes (n = 25).

| Case | Age | Sex | Primary Diagnosis | Catatonia Subtype |
| --- | --- | --- | --- | --- |
| 1 | 50s | F | Schizoaffective disorder | Malignant catatonia |
| 2 | 50s | F | Schizophrenia | Retarded catatonia |
| 3 | 50s | M | Schizophrenia | Retarded catatonia |
| 4 | 60s | M | Schizophrenia | Excited catatonia |
| 5 | 60s | M | Schizoaffective disorder | Excited catatonia |
| 6 | 60s | F | Schizoaffective disorder | Malignant catatonia |
| 7 | 40s | M | Schizophrenia | Malignant catatonia |
| 8 | 20s | F | Schizophrenia | Excited catatonia |
| 9 | 80s | M | Schizophrenia | Excited catatonia |
| 10 | 50s | F | Schizoaffective disorder | Excited catatonia |
| 11 | 40s | F | Schizophrenia | Retarded catatonia |
| 12 | 40s | F | Schizophrenia | Excited catatonia |
| 13 | 60s | F | Schizophrenia | Malignant catatonia |
| 14 | 70s | F | Schizophrenia | Excited catatonia |
| 15 | 30s | M | Schizophrenia | Excited catatonia |
| 16 | 40s | M | Schizophrenia | Excited catatonia |
| 17 | 70s | M | Schizophrenia | Retarded catatonia |
| 18 | 50s | M | Schizophrenia | Retarded catatonia |
| 19 | 70s | M | Schizophrenia | Retarded catatonia |
| 20 | 20s | F | Schizophrenia | Retarded catatonia |
| 21 | 50s | F | Schizophrenia | Retarded catatonia |
| 22 | 60s | M | Schizophrenia | Excited catatonia |
| 23 | 50s | M | Bipolar disorder | Retarded catatonia |
| 24 | 60s | M | Bipolar disorder | Excited catatonia |
| 25 | 50s | F | Major depressive disorder | Retarded catatonia |

**Supplementary Table 2.** Clinical interventions temporally associated with recovery milestones across cases (n = 25).

| Case | | Sensory-instability reduction | | Organized action | | Motivational recovery | Contextual stabilization |
| --- | --- | --- | --- | --- | --- | --- | --- |
| 1 | | Lorazepam | | Guanfacine | | – | Olanzapine |
| 2 | | Lorazepam | | Guanfacine | | – | Olanzapine |
| 3 | | Ethyl loflazepate | | Guanfacine | | – | Paliperidone LAI |
| 4 | | Lorazepam | | Guanfacine | | – | Brexpiprazole |
| 5 | | Lorazepam | | Guanfacine | | – | Brexpiprazole |
| 6 | | Pregabalin and lorazepam | | Guanfacine | | Clomipramine, lurasidone | Brexpiprazole, lurasidone |
| 7 | | Lorazepam | | Guanfacine | | – | Olanzapine |
| 8 | | Lorazepam | | Guanfacine | | – | Brexpiprazole, blonanserin |
| 9 | | Lorazepam | | Guanfacine | | – | Paliperidone |
| 10 | | Lorazepam, pregabalin | | Guanfacine | | Vortioxetine | Olanzapine, aripiprazole, valproate |
| 11 | | Lorazepam | | Guanfacine | | Vortioxetine, mirtazapine | Olanzapine |
| 12 | | Lorazepam, pregabalin | | Guanfacine | | – | Olanzapine, aripiprazole, valproate |
| 13 | | Lorazepam | | Guanfacine | | – | Olanzapine |
| 14 | | Lorazepam | | Lorazepam | | – | Paliperidone |
| 15 | | Lorazepam | | Lorazepam | | – | Quetiapine |
| 16 | | Lorazepam | | Lorazepam | | – | Quetiapine |
| 17 | | Diazepam | | Diazepam | | – | Olanzapine, lurasidone |
| 18 | | Lorazepam | | Lorazepam | | – | Quetiapine |
| 19 | | Lorazepam | | Lorazepam | | Mirtazapine | Quetiapine |
| 20 | | Lorazepam | | Lorazepam | | – | Aripiprazole |
| 21 | | Lorazepam | | Lorazepam | | – | Clozapine, valproate |
| 22 | | Lorazepam, memantine | | Guanfacine | | – | Olanzapine, brexpiprazole, memantine |
| 23 | | Lorazepam, memantine, pregabalin | | Guanfacine | | Lurasidone, olanzapine | Lurasidone, olanzapine, memantine |
| 24 | | Lorazepam | | Guanfacine | | Aripiprazole, lurasidone | Aripiprazole, lurasidone |
| 25 | | Lorazepam | | Lorazepam | | Olanzapine | Olanzapine |

*Entries indicate clinical interventions in use around the time of each recovery milestone. Temporal association does not imply a specific pharmacological effect on the corresponding computational parameter.*

**Supplementary Table 3. Bush–Francis Catatonia Rating Scale total before and after intervention (n = 25).**

| Case | Peak BFCRS total | BFCRS total after remission |
| --- | --- | --- |
| 1 | 34 | 0 |
| 2 | 14 | 0 |
| 3 | 18 | 3 |
| 4 | 28 | 3 |
| 5 | 34 | 4 |
| 6 | 28 | 2 |
| 7 | 28 | 6 |
| 8 | 19 | 0 |
| 9 | 25 | 2 |
| 10 | 34 | 0 |
| 11 | 30 | 5 |
| 12 | 29 | 4 |
| 13 | 24 | 5 |
| 14 | 33 | 0 |
| 15 | 32 | 0 |
| 16 | 26 | 0 |
| 17 | 28 | 5 |
| 18 | 21 | 0 |
| 19 | 18 | 4 |
| 20 | 26 | 0 |
| 21 | 26 | 0 |
| 22 | 25 | 0 |
| 23 | 33 | 7 |
| 24 | 23 | 0 |
| 25 | 22 | 0 |
| Mean ± s.d. | 26.3 ± 5.6 | 2.0 ± 2.4 |

Wilcoxon signed-rank test, z = 4.38, *P* = 1.2 × 10⁻⁵, r = 0.88. Scored retrospectively from records (23 items, 0–3). Elicited signs requiring a specific bedside manoeuvre were frequently not performed and, where not documented, were recorded as absent; item totals therefore reflect documented findings and may underestimate these signs. Because documentation tends to be denser at acute peak than during remission, differential ascertainment may also overstate the reduction. In a pre-specified sensitivity analysis limited to the 16 signs scorable by observation without a specific elicitation manoeuvre, scores fell from 22.5 ± 6.1 to 1.9 ± 2.2 (Wilcoxon z = 4.38, *P* = 1.2 × 10⁻⁵, r = 0.88), decreasing in all 25 patients; the excluded manoeuvre-dependent items (rigidity, waxy flexibility, gegenhalten, mitgehen, ambitendency, grasp reflex, automatic obedience) contributed 14% of the peak score.

**Supplementary Table 4. Prevalence of individual Bush–Francis Catatonia Rating Scale items at peak (n = 25).**

| Item | Present (score ≥ 1), n (%) | Score 1, n | Score 2, n | Score 3, n |
| --- | --- | --- | --- | --- |
| 1. Excitement | 16 (64%) | 2 | 1 | 13 |
| 2. Stupor | 15 (60%) | 3 | 8 | 4 |
| 3. Mutism | 18 (72%) | 0 | 8 | 10 |
| 4. Staring | 17 (68%) | 9 | 4 | 4 |
| 5. Posturing / Catalepsy | 18 (72%) | 5 | 7 | 6 |
| 6. Grimacing | 19 (76%) | 3 | 12 | 4 |
| 7. Echophenomena | 9 (36%) | 1 | 6 | 2 |
| 8. Stereotypy | 14 (56%) | 3 | 7 | 4 |
| 9. Mannerisms | 14 (56%) | 0 | 8 | 6 |
| 10. Verbigeration | 17 (68%) | 4 | 8 | 5 |
| 11. Rigidity† | 19 (76%) | 5 | 4 | 10 |
| 12. Negativism | 24 (96%) | 4 | 9 | 11 |
| 13. Waxy flexibility† | 14 (56%) | 4 | 6 | 4 |
| 14. Withdrawal | 16 (64%) | 1 | 7 | 8 |
| 15. Impulsivity | 14 (56%) | 1 | 2 | 11 |
| 16. Automatic obedience† | 1 (4%) | 0 | 0 | 1 |
| 17. Mitgehen† | 0 (0%) | 0 | 0 | 0 |
| 18. Gegenhalten† | 6 (24%) | 2 | 4 | 0 |
| 19. Ambitendency† | 4 (16%) | 2 | 0 | 2 |
| 20. Grasp reflex† | 2 (8%) | 1 | 1 | 0 |
| 21. Perseveration | 14 (56%) | 2 | 9 | 3 |
| 22. Combativeness | 15 (60%) | 3 | 3 | 9 |
| 23. Autonomic abnormality | 13 (52%) | 4 | 7 | 2 |

Scored retrospectively from records at the time of peak severity, as in Supplementary Table 3. A score of 1 or more is counted as present. † Elicited sign requiring a specific bedside manoeuvre; where the manoeuvre was not documented the item was recorded as absent, so these counts may underestimate prevalence. Every patient showed at least 7 of the 23 items (median 12, range 7–15). Negativism was the most frequent item (24 of 25); mitgehen was not documented in any patient.

**Supplementary Table 5.** Recovery milestone timing and its relation to baseline severity (n = 25). Day counted from symptom onset (day of onset = day 1). ρ is the Spearman rank correlation between the peak (pre-treatment) BFCRS total score and the day on which the milestone was reached; for πv_slow this is also the interval from symptom onset to the final milestone. Peak BFCRS total: median 26 (IQR 23–30), range 14–34. These correlations are exploratory and the *P* values are not corrected for multiple comparisons. The three πv_fast observations recorded as not applicable were coded not assessable by the primary rater: an antipsychotic had been given from initiation and the baseline record was ambiguous as to whether the disturbance the criterion requires had been present, so no date was assigned. The blind rater dated πv_fast in all three.

| **Domain** | **n applicable** | **Median day (IQR)** | **Range** | **ρ with peak BFCRS (*P*)** |
| --- | --- | --- | --- | --- |
| πs | 25 | 3 (2–7) | 1–18 | 0.13 (0.54) |
| β | 25 | 4 (3–11) | 1–21 | 0.11 (0.59) |
| πm | 25 | 8 (3–14) | 1–21 | 0.22 (0.29) |
| πv_fast | 22 | 14.5 (6.8–23.2) | 1–40 | 0.38 (0.08) |
| πv_slow | 25 | 15 (8–27) | 2–42 | 0.29 (0.16) |

**Supplementary Table 6.** Adjacent-pair ordering of recovery milestones. Predicted earlier → later domain. Consistent = earlier ≤ later (same-day ties included). Sign test on non-tied pairs. Separable pairs (πs → β; πm → πv_fast), whose definitions do not presuppose one another, show that the observed order was not entailed by the definitions (though not that the domains are mechanistically independent); nested pairs (β → πm; πv_fast → πv_slow) share a definitional dependency in rank order, for which the empirical quantity is inter-milestone latency.

| Pair | n | Strict precedence | Same day | Inversion | Consistent (≤) | Sign-test *P* |
| --- | --- | --- | --- | --- | --- | --- |
| πs → β | 25 | 15 | 10 | 0 | 25/25 (100%) | <0.001 |
| β → πm | 25 | 12 | 13 | 0 | 25/25 (100%) | <0.001 |
| πm → πv_fast | 22 | 18 | 4 | 0 | 22/22 (100%) | <0.001 |
| πv_fast → πv_slow | 22 | 22 | 0 | 0 | 22/22 (100%) | <0.001 |

**Supplementary Table 7.** Blinded re-rating of milestone dates (I.M.): adjacent-pair ordering in the blind ratings (a) and agreement with the primary coding by domain (b). Pre-registered plan: Zenodo doi:10.5281/zenodo.23076104; analysis script: doi:10.5281/zenodo.23388109. No value in this table was known when the plan was deposited.

**a. Adjacent-pair ordering in the blind ratings.** Predicted earlier → later domain. n = patients in whom both milestones carried a date in the blind ratings. Separable pairs are the primary criterion: any inversion refutes the pre-specified order. Sign test two-sided, on non-tied pairs.

| Pair | Separable | n | Strict precedence | Same day | Inversion | Sign-test P |
| --- | --- | --- | --- | --- | --- | --- |
| πs → β | Yes | 25 | 16 | 9 | 0 | <0.001 |
| β → πm | No (nested) | 25 | 11 | 14 | 0 | <0.001 |
| πm → πv_fast | Yes | 25 | 17 | 8 | 0 | <0.001 |
| πv_fast → πv_slow | No (nested) | 25 | 24 | 1 | 0 | <0.001 |

**b. Agreement with the primary coding by domain.** Day counted from symptom onset (day of onset = day 1). ICC(A,1): two-way, absolute agreement, single rater, 95% confidence interval. Bland–Altman: mean of (blind minus primary) with 95% limits of agreement. Applicability: a date in both ratings versus NA or ND in either; Cohen’s κ for πv_fast, the only domain with a non-dated entry in either rating. Kendall’s W (tie-corrected) over patients with five dates in the blind ratings; primary value 0.91 (n = 22).

| Domain | Primary: dated / NA | Blind: dated / NA / ND | ICC(A,1) (95% CI) | Bland–Altman mean (LoA), days | Median \|difference\|, days |
| --- | --- | --- | --- | --- | --- |
| πs | 25 / 0 | 25 / 0 / 0 | 0.71 (0.46 to 0.86) | 1.00 (−5.67 to 7.67) | 1 |
| β | 25 / 0 | 25 / 0 / 0 | 0.72 (0.42 to 0.87) | 2.12 (−5.91 to 10.15) | 2 |
| πm | 25 / 0 | 25 / 0 / 0 | 0.75 (0.50 to 0.88) | 1.96 (−7.41 to 11.33) | 3 |
| πv_fast | 22 / 3 | 25 / 0 / 0 | 0.90 (0.76 to 0.96) | −1.55 (−10.71 to 7.62) | 2 |
| πv_slow | 25 / 0 | 25 / 0 / 0 | 0.84 (0.66 to 0.92) | 1.04 (−13.96 to 16.04) | 1 |
| Applicability, κ (95% CI) |  |  | 0 (πv_fast; blind ratings contained no NA or ND) |  |  |
| Within-patient order, median τ-b (range) |  |  | 0.95 (0.67 to 1.00); n = 25 |  |  |
| Kendall’s W, blind (n) |  |  | 0.89 (n = 25) |  |  |

**Supplementary Table 8.** Primary and blind milestone days for every patient. Day counted from symptom onset (day of onset = day 1); primary / blind. Pairs in which the two ratings differ (different day, or NA in the primary coding against a date in the blind rating) are marked with an asterisk; 87 of 125 patient-domain pairs differ. The blind sheet recorded days only, so the record entries behind each rating were not collected. Discrepancies were examined only after the ratings were locked; neither rating was altered.

| Case | πs | β | πm | πv_fast | πv_slow | Discrepant domains |
| --- | --- | --- | --- | --- | --- | --- |
| 1 | 6 / 13* | 7 / 14* | 14 / 17* | 21 / 17* | 23 / 23 | πs, β, πm, πv_fast |
| 2 | 7 / 7 | 14 / 14 | 14 / 24* | NA / 64* | 31 / 64* | πm, πv_fast, πv_slow |
| 3 | 2 / 2 | 3 / 3 | 3 / 3 | NA / 3* | 5 / 6* | πv_fast, πv_slow |
| 4 | 3 / 4* | 4 / 17* | 6 / 18* | 19 / 19 | 21 / 22* | πs, β, πm, πv_slow |
| 5 | 2 / 3* | 3 / 4* | 4 / 4 | 9 / 18* | 14 / 20* | πs, β, πv_fast, πv_slow |
| 6 | 1 / 1 | 4 / 1* | 4 / 1* | 5 / 1* | 7 / 2* | β, πm, πv_fast, πv_slow |
| 7 | 7 / 10* | 11 / 11 | 15 / 27* | 28 / 28 | 31 / 30* | πs, πm, πv_slow |
| 8 | 3 / 3 | 3 / 5* | 3 / 7* | 13 / 14* | 14 / 15* | β, πm, πv_fast, πv_slow |
| 9 | 8 / 9* | 10 / 14* | 14 / 17* | 17 / 21* | 22 / 24* | πs, β, πm, πv_fast, πv_slow |
| 10 | 18 / 19* | 21 / 24* | 21 / 24* | 40 / 38* | 42 / 49* | πs, β, πm, πv_fast, πv_slow |
| 11 | 2 / 2 | 2 / 4* | 8 / 10* | 14 / 10* | 15 / 16* | β, πm, πv_fast, πv_slow |
| 12 | 10 / 12* | 11 / 12* | 11 / 12* | 34 / 30* | 35 / 32* | πs, β, πm, πv_fast, πv_slow |
| 13 | 11 / 4* | 11 / 18* | 11 / 18* | 19 / 19 | 20 / 20 | πs, β, πm |
| 14 | 6 / 6 | 11 / 15* | 14 / 15* | 29 / 23* | 30 / 31* | β, πm, πv_fast, πv_slow |
| 15 | 2 / 2 | 2 / 2 | 3 / 3 | 4 / 3* | 6 / 4* | πv_fast, πv_slow |
| 16 | 3 / 2* | 12 / 8* | 12 / 8* | 12 / 10* | 14 / 15* | πs, β, πm, πv_fast, πv_slow |
| 17 | 1 / 1 | 1 / 1 | 1 / 1 | NA / 1* | 2 / 2 | πv_fast |
| 18 | 4 / 14* | 6 / 15* | 17 / 20* | 27 / 24* | 28 / 25* | πs, β, πm, πv_fast, πv_slow |
| 19 | 1 / 1 | 1 / 1 | 1 / 1 | 1 / 1 | 4 / 2* | πv_slow |
| 20 | 2 / 2 | 2 / 2 | 2 / 2 | 6 / 4* | 8 / 5* | πv_fast, πv_slow |
| 21 | 3 / 2* | 3 / 3 | 14 / 4* | 24 / 7* | 27 / 14* | πs, πm, πv_fast, πv_slow |
| 22 | 9 / 7* | 10 / 7* | 10 / 10 | 11 / 12* | 12 / 13* | πs, β, πv_fast, πv_slow |
| 23 | 4 / 13* | 4 / 13* | 8 / 13* | 15 / 14* | 16 / 20* | πs, β, πm, πv_fast, πv_slow |
| 24 | 2 / 3* | 4 / 5* | 5 / 5 | 5 / 6* | 8 / 7* | πs, β, πv_fast, πv_slow |
| 25 | 2 / 2 | 2 / 2 | 2 / 2 | 2 / 2 | 3 / 3 | none |

Supplementary Note 1. Retrospective labelling of agents by precision domain

Pharmacological interventions were selected by the treating physicians on clinical grounds during routine care. No protocol derived from the framework was applied, and no study measure was added to routine care. Dosing and sequencing were individualized by the treating physicians according to symptom severity, medical risk, prior treatment response and overall clinical judgement. The scheme below is a retrospective interpretive labelling of the record, imposed by the first author (HS) after the fact and solely for correspondence with Supplementary Table 2; it formed no part of clinical decision-making, and no agent was chosen with any of these labels in view. Agents already in use during the early period, when autonomic instability and exaggerated reactivity predominated, are labelled here in relation to sensory precision (πs). α2A-adrenergic modulation, used in a subset of the cohort on clinical grounds unrelated to the present scheme, happened in those cases to be in use around the time organized action stabilized, and is labelled in relation to policy precision (β). Agents in use around the return of spontaneous initiation are labelled in relation to motivational precision (πm), and agents in use around the containment of short-timescale instability in relation to fast contextual volatility (πv_fast). No agent in this cohort falls under the label of slow structural volatility (πv_slow); that domain changed over the same period with no agent to which it could be assigned. The labels therefore record which agents happened to be in use when a given milestone was reached. As in Supplementary Table 2, they denote temporal co-occurrence only, and do not imply that any agent acted on the corresponding computational parameter, nor that the correspondence was noticed at the time. The experimental basis for the α2A-adrenoceptor rationale, and its limits, are given in the Methods of the main text; it is cited there as background, not as evidence that any agent acted on the corresponding domain.

Supplementary Note 2. Aetiological scope, and the pharmacology of memantine in relation to NMDA receptor autoantibodies

The cohort reported here was assembled at a psychiatric hospital without electroconvulsive therapy capability and is accordingly weighted towards catatonia arising in psychotic and affective disorders (Supplementary Table 1). Catatonia also arises from epileptic, hypoxic, metabolic, infectious and autoimmune causes.^1,2^ Hospital-wide, about one fifth of catatonia has a medical cause, the proportion exceeds one half in acute medical and surgical settings, and central nervous system disease accounts for roughly two thirds of those cases.^1^ The autoimmune encephalitides, in particular those associated with antibodies to the N-methyl-D-aspartate (NMDA) receptor, are of specific interest here, because catatonic features are frequent in that syndrome and because first-episode psychosis cohorts have been screened for the same antibodies.^3,4^

The single patient with major depressive disorder (case 25) is not separated from the cohort in the milestone data. Sensory precision, policy precision, motivational precision and fast contextual volatility were reached on day 2 and slow structural volatility on day 3, so the case is early rather than aberrant: it is the minimum in no domain, ranks second to fourth among the 22 to 25 patients contributing each milestone, and no value lies beyond 1.5 interquartile ranges from the quartiles of its domain (Supplementary Table 5). Because its first four milestones fell on the same day, it carries order information for the final adjacent pair only, and it contributes no inversion at any pair (Supplementary Table 6). The two patients with bipolar disorder (cases 23 and 24) likewise fall within the cohort distributions.

The recovery ordering does not reference aetiology. It follows from two conditions on the recovery dynamics: a common impaired onset across the five domains, and a drive that is positive in the residual deficit and vanishes as that deficit closes. Both are where a secondary cause is expected to depart from the presentation studied here. An insult that is focal, as in epilepsy, regionally selective, as in hypoxic-ischaemic injury, or receptor-selective, as in antibody-mediated encephalitis, need not impair the five domains uniformly, so the onset is not common across domains. This does not abolish the prediction. Non-uniform onset permits only a restricted set of inversions, each at the adjacent pair straddling the domain the illness spared, and the companion theoretical work names which inversion a given onset profile permits.

The second condition fails for a different reason. While the cause is still active, with antibodies still circulating, a metabolic derangement uncorrected or seizures uncontrolled, a process opposing recovery runs concurrently with the deficit-closing drive, so the drive is not of the assumed form. The prediction is therefore specific and dated: the ordering should hold once the cause has been removed, that is, after immunotherapy has taken effect, after the metabolic derangement has been corrected or after seizures have been controlled, and should not be expected to hold while that removal is in progress. Testing this requires cohorts with an established aetiology and documented milestone times, which the present series does not contain.

Memantine was used adjunctively in two cases (Supplementary Table 2). Because it is an NMDA receptor antagonist, it might be expected to reproduce rather than relieve the state associated with NMDA receptor autoantibodies, which is frequently accompanied by catatonic features. The two act on the receptor in different ways. Patient antibodies against the GluN1 subunit bind, cap and cross-link surface receptors and drive their internalisation, giving a selective and reversible fall in receptor surface density and synaptic localisation that tracks antibody titre; synaptic NMDA receptor-mediated currents decrease while other receptors, synapse number and dendritic structure are spared.^5^ Memantine does not remove receptors. It is a low-affinity, uncompetitive open-channel blocker with a fast off-rate, entering the channel preferentially when it is open for prolonged periods and leaving it quickly, so that, in experimental preparations at concentrations in the therapeutic range, it attenuates tonic, largely extrasynaptic receptor activity while relatively sparing the millisecond-scale phasic transmission that carries synaptic signalling.^6,7^ Whether this relative sparing holds under clinical conditions has not been shown.

The distinction is between removing receptors available for phasic synaptic transmission and lowering the tonic component of activity while that transmission is relatively preserved. In the terms used here, the antibody removes the channel through which sensory evidence reaches higher levels of inference, whereas memantine reduces the gain on that evidence, which is the direction in which excessive sensory precision (πs) is attenuated without disabling the inference that depends on it. This is consistent with where memantine appears in Supplementary Table 2, in the sensory-instability and contextual-stabilization columns rather than at the motivational milestone.

This is an interpretation of an agent selected on clinical grounds in two cases, not a controlled comparison, and, as in Supplementary Note 1, the grouping of interventions by precision domain denotes temporal association only.

**Supplementary Note 3. Worked examples of milestone coding**

The following are representative record entries and the assignments they produced. They are given so that the amount of judgement involved can be seen, and so that an independent rater can apply the same definitions. Entries are paraphrased and de-identified.

Sensory precision (πs). Entry: 'Startles and pulls away when the curtain is drawn; heart rate 120, sweating; settles after the room is darkened.' Coded as baseline disturbance present. The milestone was dated from a later entry, 'tolerates ward noise, no autonomic surge during care, sleeps four hours uninterrupted', which positively documents the capacity rather than merely omitting the abnormality.

Policy precision (β). Entry: 'Does not move spontaneously. When the spoon is brought to the mouth, opens and swallows; each mouthful requires a fresh prompt.' Coded as the β milestone, not πm, because every step required an external prompt. Ambiguity rule applied: where the record does not state whether the behaviour was prompted, the conservative assignment (β) was taken.

Motivational precision (πm). Entry: 'Picked up chopsticks and ate without being asked; asked the nurse for tea.' Coded as the πm milestone, because initiation was unprompted and continued beyond a single prompt. An entry reading 'cooperative with feeding' would not have been sufficient.

Fast contextual volatility precision (πv_fast). Entry: 'Repeats the examiner's last words; switches abruptly from conversation to laughing; responsiveness fluctuates within the same interview.' Coded as baseline disturbance present. The milestone was dated from the first sustained interval, within a day or across repeated interactions within a day, documented free of such short-timescale instability. In three patients an antipsychotic was given from initiation and the baseline record was ambiguous as to whether such instability had been present; as the conservative choice, these were coded not assessable rather than dated.

Slow structural volatility precision (πv_slow). Entry: 'Slept through the night; same level of engagement with the day and night nursing staff; account of why she is in hospital is consistent across two interviews.' Coded as the πv_slow milestone. This domain corresponds to no single DSM-5-TR criterion and was ascertained only from entries of this kind.

Multi-determined signs. Agitation is compatible with elevated sensory precision, reduced policy precision and elevated fast contextual volatility. It was not assigned to a mechanism; the entry was used only where it satisfied a specific milestone definition, and the alternative assignment was recorded. Reassigning such a sign changes which behaviour indexes a milestone, not the predicted order of domain stabilisation.

Supplementary Note 4. Blinded re-rating of milestone dates: procedure and label correspondence

After the primary analysis had been completed, the five milestone dates were re-rated for all 25 patients from the medical records by a psychiatrist at the same hospital (I.M.) who had taken no part in the study and had not seen the manuscript, the preprint or the companion theoretical papers. The rater was not told the hypothesised order, the first rater’s dates, or the correspondence between the labels and the precision domains. The rater received the milestone definitions given in the Methods, labelled A to E in an order unrelated to the hypothesised sequence, a short rating manual in Japanese, and a rating sheet with one row per patient and one column per label. The label correspondence, fixed before the manual was handed over and deposited with the plan, was A = πv_fast, B = πv_slow, C = πm, D = πs, E = β.

Permitted entries were a calendar date (the first day on which the definition was met and the record showed it to be sustained), NA (the baseline disturbance the definition requires was not documented), or ND (the baseline disturbance was documented but no entry recording its resolution was found before discharge). The manual instructed the rater, where two days were equally defensible, to take the later day, and where it was doubtful whether the definition had been met, to treat it as not yet met; both rules err towards dating milestones late, as in the primary coding. No confidence rating was collected, no second pass was permitted, and the sheet was locked on return. The returned sheet gave each milestone as a day count from the reference date of the primary analysis (day of onset = day 1); every cell carried a day, and no NA or ND entry was returned.

The analysis plan (version 1.0, dated 1 October 2026), the manual and the label correspondence were deposited at Zenodo (doi:10.5281/zenodo.23076104) before any rating had been returned, and the deposit was disclosed by e-mail to M.T. on the same day; it was held under embargo while the ratings were collected and was made open on 16 October 2026. The analysis script (blind_rerating_analysis.py) was deposited on 15 October 2026 with the figure code (doi:10.5281/zenodo.23388109). The public metadata of the deposit do not state the hypothesised order. The plan fixed as the primary criterion the number of inversions of the two separable transitions (πs → β; πm → πv_fast) in the blind ratings, with a single inversion refuting the pre-specified order; the nested pairs are reported descriptively; Kendall’s W is reported beside the primary value without a threshold; and date agreement, applicability agreement and within-patient order agreement are summarised by ICC(A,1), Cohen’s κ and Kendall’s τ-b respectively. Analyses were run in IBM SPSS Statistics v32 and reproduced with the deposited script. The rater was unblinded, and joined the author list, only after the ratings had been locked. The returned sheet carries no calendar information; both raters’ day counts from onset are given in Supplementary Table 8 and in the public code repository, and the medical records behind the ratings are not public.
